# COVID-19 Vaccine hesitancy in Addis Ababa, Ethiopia: A mixed-methods study

**DOI:** 10.1101/2021.02.25.21252443

**Authors:** Nebiyu Dereje, Abigel Tesfaye, Beamlak Tamene, Dina Alemeshet, Haymanot Abe, Nathnael Tesfa, Saron Gedion, Tigist Biruk, Yabets Lakew

## Abstract

**Background:** COVID-19 infection is a global pandemic threatening the public health. Due to the development and initiation of vaccination, currently significant difference upon vaccine acceptance is seen between developed and developing countries. However, there are no data on the level of COVID-19 vaccine hesitancy and its associated factors in Addis Ababa, Ethiopia.

**Methods:** An embedded mixed method study [QUAN(quali)] was conducted among residents of Akaki Kality sub-city in Addis Ababa, Ethiopia. For the quantitative part, a multi-stage sampling technique was used to recruit the study participants (n = 422). Twenty four adults were included purposively for the qualitative in-depth interview. Data was collected by face-to-face interview by using a semi-structured questionnaire. Factors associated with COVID-19 vaccine hesitancy were identified by multivariable binary logistic regression model.

**Result:** The mean age of the participants was 34.1 years (±12.9). Nearly half (46.7%) of the participants exhibited poor level of knowledge and 51.8% had negative attitude towards COVID-19 and its preventive measures. One out five (19.1%) participants were not willing to get vaccinated when it becomes available. In the multivariable analysis, vaccine hesitancy was significantly associated with being female (aOR=1.97; 95% CI: 1.10 - 3.89, p=0.03), negative attitude towards COVID-19 and its preventive measures (aOR=1.75; 95% CI: 1.08 - 3.02, p=0.04), and information source being social media (internet) (aOR=3.59; 95% CI: 1.75 - 7.37, P <0.0001).

**Conclusion:** A considerable proportion of the people in Addis Ababa have concerns on COVID-19 vaccine and unwilling to accept once it is available. Several conspiracy theories were put forth to justify their stance and this was mainly due to the misconceptions distributed from the use of social media as source of information. Overall, providing the community with health education and consistent government efforts in uphold the prevention measures are of paramount importance to tackle this pandemic.

## Introduction

Corona virus disease 2019 (COVID-19) is caused by severe acute respiratory syndrome corona virus 2 (SARS-CoV-2) also known as Novel coronavirus (nCov) [1]. The first case of COVID-19 was discovered in Wuhan city, Hubei province of China with unexplained pneumonia on December 12, 2019 [2]. The virus is transmitted through large droplets generated during coughing or sneezing of symptomatic and asymptomatic patients [3]. Therefore, frequent hand-washing with soap and water, using alcohol based hand rub or sanitizer, avoidance of hand shaking/public gathering and use of face mask are crucial to halt the spread of COVID-19 [4]. COVID-19 was declared a pandemic by the World health organization on March 11, 2020 [5]. Since its emergence, this pandemic has shown its capability to spread rapidly in the world causing the most dramatic global health crisis of our time resulting in devastating social, economic and political crises [6].

Globally, more than 210 countries/territories have been affected by the virus, with more than one hundred ten million people being infected and 2.5 million deaths reported as of February 24, 2021. Ethiopia ranks 71^st^ regarding COVID-19 with more than 154, 000 infected and 2,305 dead (February 24, 2021) [7]. Unfortunately, Ethiopia was found to be one of five African countries with the highest case burden of COVID-19 [7]. However, the government of Ethiopia has been striving to spread information on COVID-19 preventive measures via television, radio or social media outlets and declared a state of emergency [8].

Currently, COVID-19 vaccine has been made available but it is highly controversial. More than seven billion doses have been pre-purchased by countries and organizations of the world, of which more than half was sold-out to high income countries [9]. This figure is threatening to the global health as may be an indication of the disparities on the health delivery globally.

Myths and conspiracy theories on vaccinations have been spreading and can easily be accepted by the developing world. This may cause people to be reluctant and maleficent towards vaccination, which has been demonstrated by a study in Nigeria by a low vaccine acceptability rate [10]. WHO defined vaccine hesitancy as it is a difficulty in accepting or an outright refusal of vaccines, despite their availability. In 2019, before the COVID-19 pandemic, the World Health Organization listed vaccine hesitancy as one of the ten global threats to public health [11]. Currently, Ethiopia has not started administering the vaccines.

Hence it is very crucial to understand the varying vaccine attitudes among the community to design a strategy to overcome the vaccine hesitancy. Furthermore, unraveling the specific fears and doubts of the community with regards to receiving the vaccine can help government and other concerned officials to adequately address the misconceptions and various conspiracy theories in their campaigns. Therefore, this study was aimed to assess the level of COVID-19 vaccine acceptability among the population in Addis Ababa, the capital city of Ethiopia by employing a community based mixed methods study.

## Methods and Materials

### Study design and participants

A concurrent embedded mixed methods design [QUAN(qual)] was employed from January 20 – 31, 2021 among adult population (≥18 years) currently residing in Akaki Kality sub city of Addis Ababa, Ethiopia. The qualitative part of the study was embedded in the bigger quantitative cross-sectional study. The qualitative part was mainly intended to explain the reasons for COVID-19 vaccine hesitancy, as a supplementary of the quantitative study.

A sample size for the quantitative part of the study (n = 422) was determined by using a single population proportion formula, by taking 95% confidence interval, 5% margin of error, 50% proportion of vaccine hesitancy and adding up 10% non-response rate. For the qualitative part, 24 participants were included into the study based on the information saturation.

Multi-stage sampling technique was employed to recruit the participants for the quantitative part. There were 13 districts in the sub-city; of which three of them were selected randomly (lottery method). The total sample was allocated proportionally to the districts. Then, the households from each district were selected by employing a systematic random sampling (sampling interval = every 4^th^ house). From the specific households selected, only one randomly selected eligible individual was interviewed. For the qualitative part of the study, purposive sampling method was used to recruit participants who have reach information.

### Data collection tools and procedures

Data was collected by using a semi-structured questionnaire which was adapted from reviewed literatures [10, 12, 13]. The questionnaire has 5 components: socio-demographic, knowledge towards COVID-19, attitude towards COVID-19, practice of COVID-19 prevention measures, and COVID-19 vaccine acceptance. The questionnaire was in English and translated into Amharic for the interview. The questionnaire was administered face-to-face by the medical interns. For the qualitative part of the study, in-depth interviews were made by the investigators.

### Data management and analysis

Data was coded and entered into SPSS-for windows version 25 for analysis. Frequency and proportions were used to summarize categorical variables, whereas mean and standard deviation were used to summarize continuous variables.

The primary outcome variable of the study was COVID-19 vaccine hesitancy which was assessed by asking a question “Will you get vaccinated if you get COVID-19 vaccine?” then the response was dichotomized as “Yes” or “No”.

Multivariable binary logistic regression analysis was carried out to identify factors associated with vaccine hesitancy, as expressed by adjusted odds ratio (aOR) along with its respective 95% confidence interval (CI). Variables with <0.25 in bivariate analysis were considered for multivariable analysis. Variables having *P* value <0.05 were considered statistically significant. Multicollinearity was assessed by the colleniarity diagnostics (Variance Inflation Factor and the tolerance test). Goodness of the model was checked by the Hosmer Lemshow goodness of fit test. The qualitative data analysis was initiated by transcription and translating of the interviews, then coded and analyzed by thematic analysis. The findings of the qualitative study were used to supplement the findings of quantitative data.

### Ethical consideration

Ethical approval of this study was obtained from the Institutional Review Board (IRB) of Myungsung Medical College. The participants of the study was informed about the purpose of the study and provided their written consent. At the end of the interview, the data collectors have provided information with regard to the COVID-19 vaccine.

## Result

### Socio-demographic characteristics

A total of 409 respondents completed the questionnaire, with a response rate of 96.9%. Majority of the participants 294 (71.9%) were females and married (62.3%) (Table 1). The mean age of the participants was 34.1 years (±12.9), ranging from 18 - 85 years.

**Table 1:**
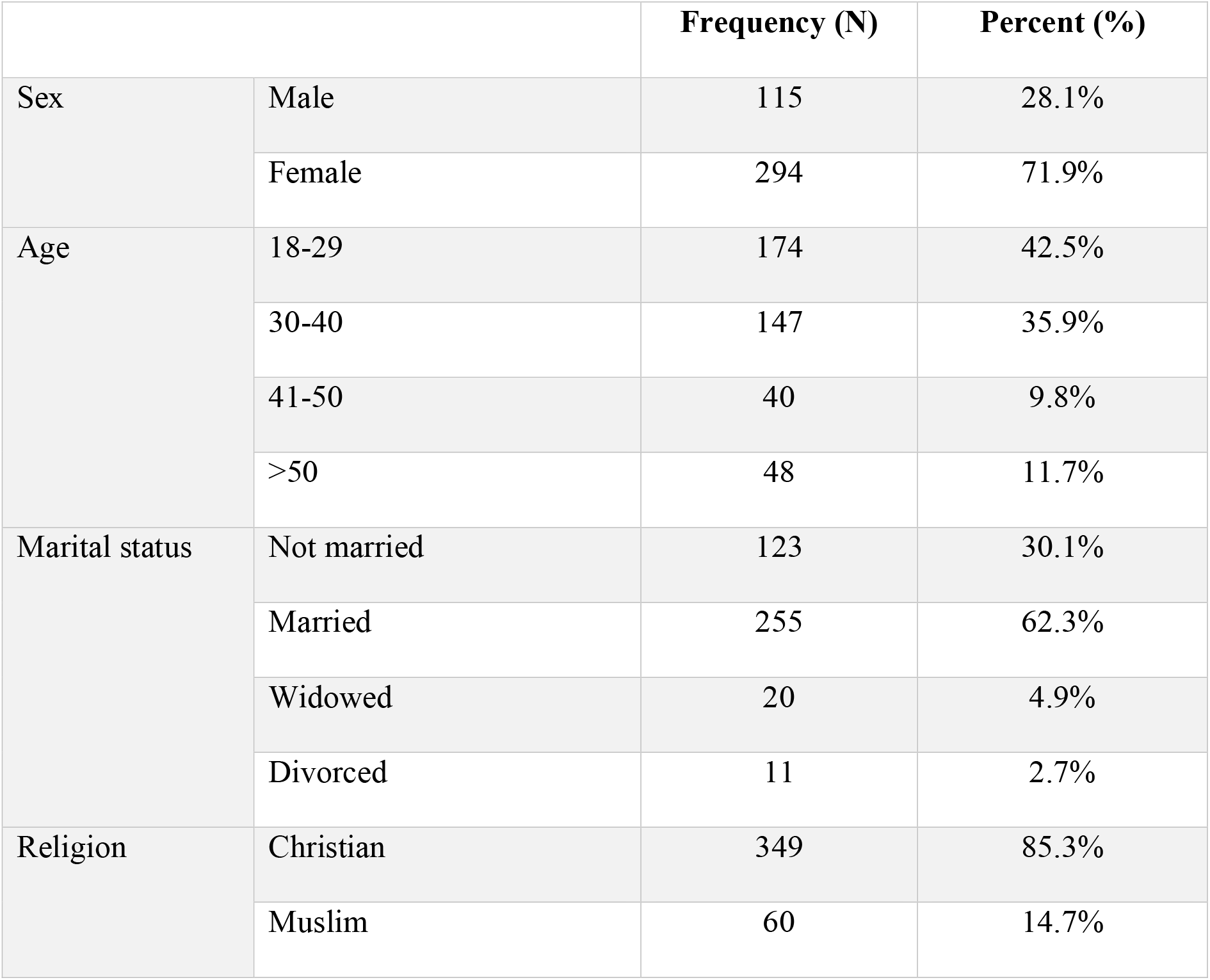
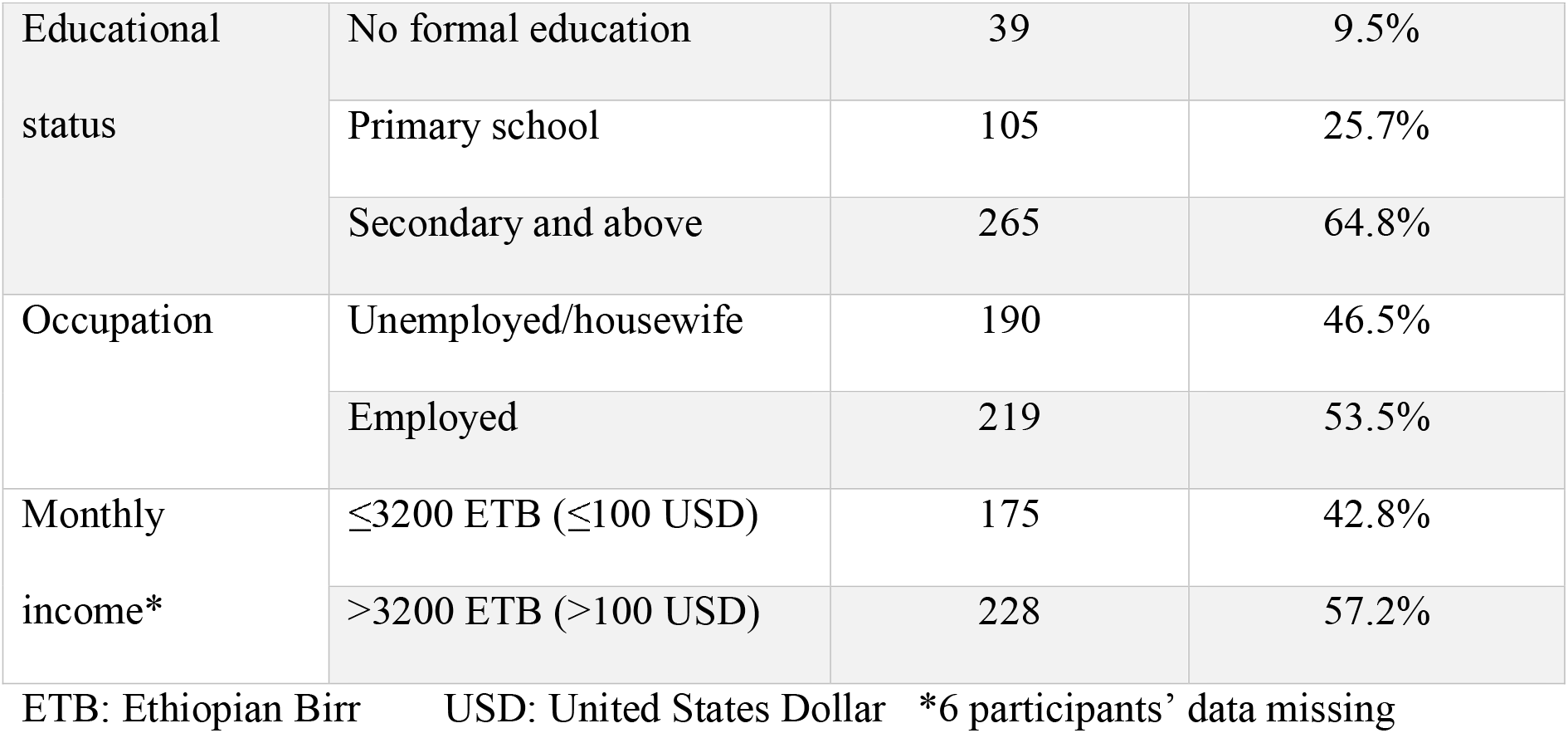
Socio-demographic characteristics of the study participants

### Knowledge towards COVID-19

Almost all of the participants heard about COVID-19 from Television or Radio. However, the average knowledge score was 56.7 ± 3.7, with 46.7% (n=191) exhibited poor level of knowledge (Supplementary Table 1).

The qualitative part of the study found that some of the study participants stated that they do not possess enough knowledge and would like to know more about things like the prospective vaccines, the new strain and how to prevent getting infected from asymptomatic patients.

For example, one 39 year old male participant said that;

> *“I want to know where the virus really came from initially; is it natural or manmade?”[39 year old, male]*

### Attitude towards COVID-19

The mean attitude score was found to be 20.3 ± 1.2, with 51.8% of the participants have negative attitude regarding COVID-19 and its preventive measures (Supplementary Table 2).

This result is corroborated by the findings on the qualitative part of the study where the majority of the respondents stated that they were initially very concerned but now they were less so. In fact, the majority of the participants stated that they were very afraid and anxious initially believing that they were definitely getting infected and that they would certainly die if they caught it. Currently, most of the respondents reported feeling less anxious. While some stated that while they were still aware that it was a serious illness they were less concerned since they believed it was preventable. While others stated the reason for their lack or decreasing concern was that they believed they would recover if they got infected.

In a spectrum of similar answers, five of the participants stood out stating that they did not believe the disease exits anymore since they have not personally encountered an infected person. In contrast to these responses, we also found two participants who stated that while they initially were not concerned they are starting to get worried now since the number of infected individuals and the resulting death toll in this country is increasing.

One female participant stated the following to show that she is not concerned about COVID-19:

> *“I am not scared because I expected this to happen; we brought this on ourselves and we are paying for our sins. It has been long time coming*.*”[Female, 50 year old]*

Another 45 year old female participant stated:

> *“I have been through an outbreak before…I got sick and I had to be isolated from my family but I recovered easily and I don’t believe this would be any different*.*” [Female, 47 year old]*

More than two-thirds (68.7%) of the participants believed Ethiopia would win the battle against the pandemic. This finding was also supported by the qualitative study.

> *“I was afraid that everyone in Ethiopia would die because even white people could not handle it. I think the only reason we have survived is because Ethiopia is God’s country*.*” [Female, 70 year old]*

### Vaccine hesitancy

More than 90% of the participants heard about the prospective COVID-19 vaccine mainly from TV/Radio. However, 78 (19.1%) were not willing to get vaccinated when it becomes available (Figure 1).

**Figure 1:**
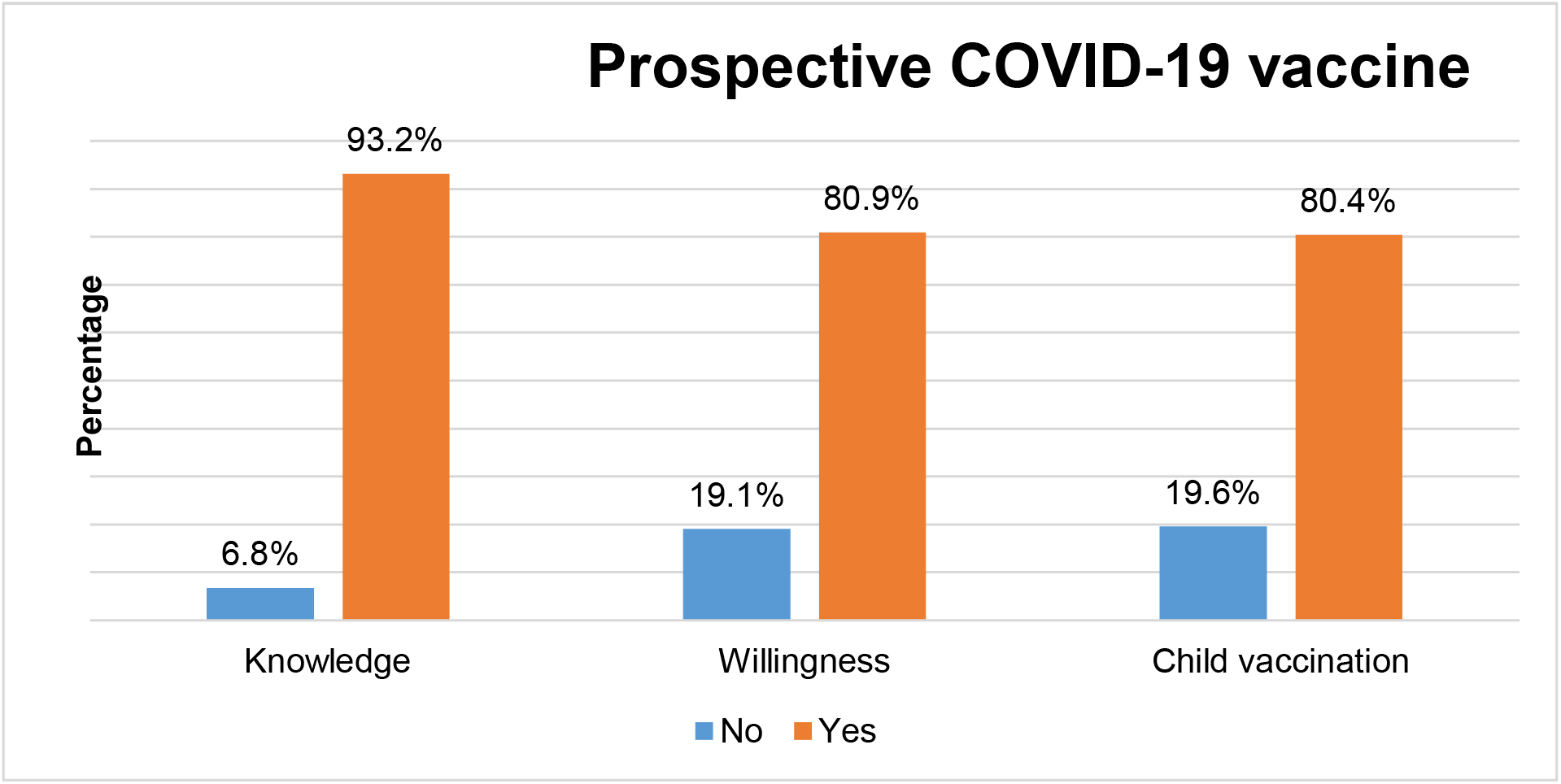
Prospective COVID-19 vaccine.

Out of them, 43.6% don’t take the vaccine due to fear of side effects and 41.0% of them believe that the vaccine may be biological weapon (Figure 2).

**Figure 2:**
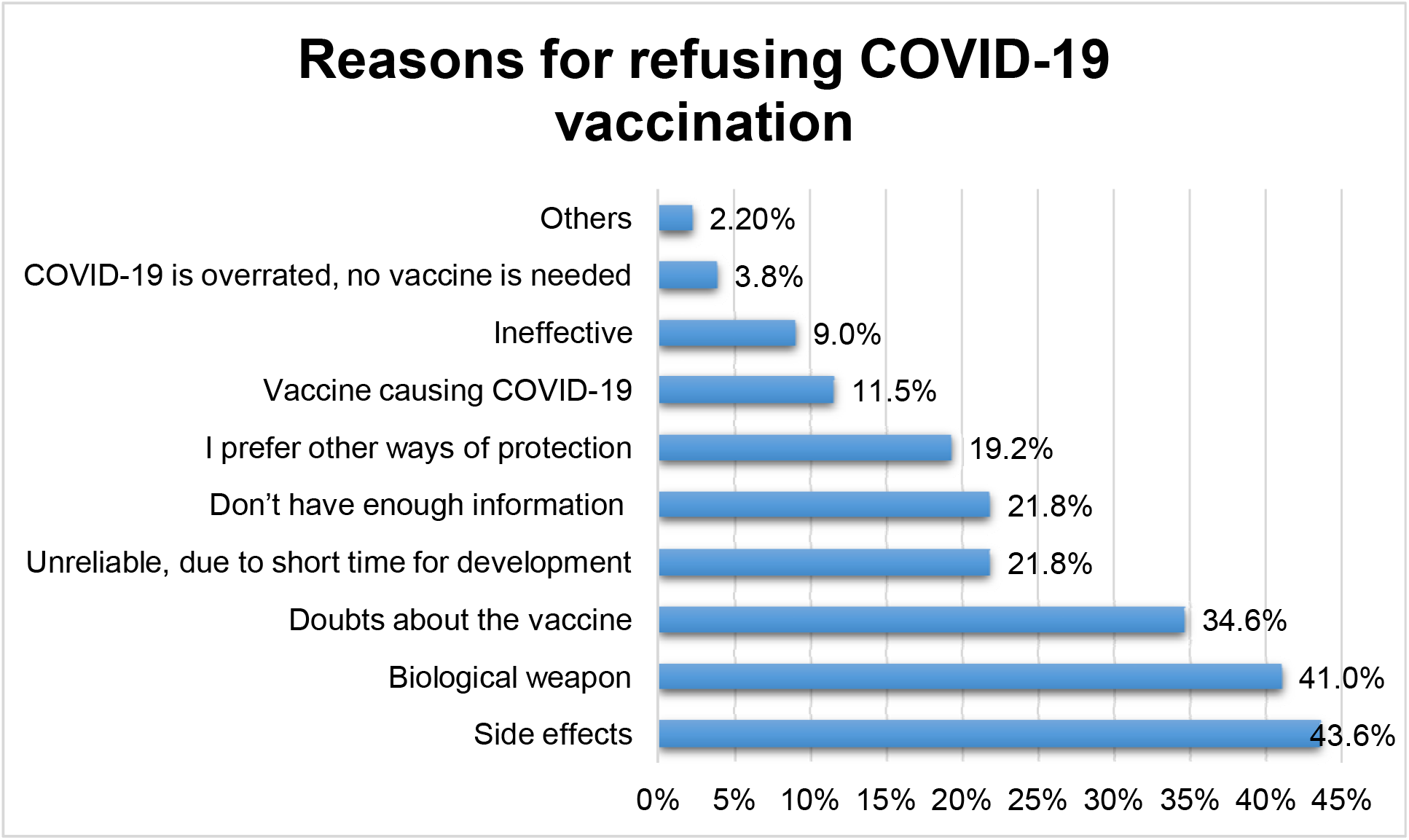
Reasons of participant for refusing COVID-19 vaccination.

In the qualitative in-depth interview, some stated they did not have enough information about the vaccine and wanted to see other people take it first. Majority of the respondents feared it would not be effective or have too many side effects as there was not enough time to study it and as a result stated that they preferred other methods to prevent COVID-19. A few of the participants thought that the vaccine that will be distributed in Africa would be of lower quality. Others thought it would be used as a biological weapon by the developed nations to cause infertility and control the population of poor countries. Moreover, it was also mentioned that the vaccine might be used as a weapon to insert microchips made by the illuminati, into the body as the mark of the beast “(666)” that would cause them to forsake their faith. A few others did not think they needed the vaccine because they had God’s protection.

> *“I don’t think the vaccine will come to this country and even if it does I don’t need it; God will be my vaccine*.*” [Female, 45 year old]*

Close to 20% of the participants thought that children should not get vaccinated. Some of the participants did not recommend the vaccine to children even though they would take it themselves. These participants further expressed in the in depth interview that they thought th virus did not affect children or it would be too dangerous for them.

### Factors associated with vaccine hesitancy

In the multi-variable analysis (Table 2), COVID-19 vaccine hesitancy was associated with sex, attitude and source of information about the vaccine.

**Table 2:**
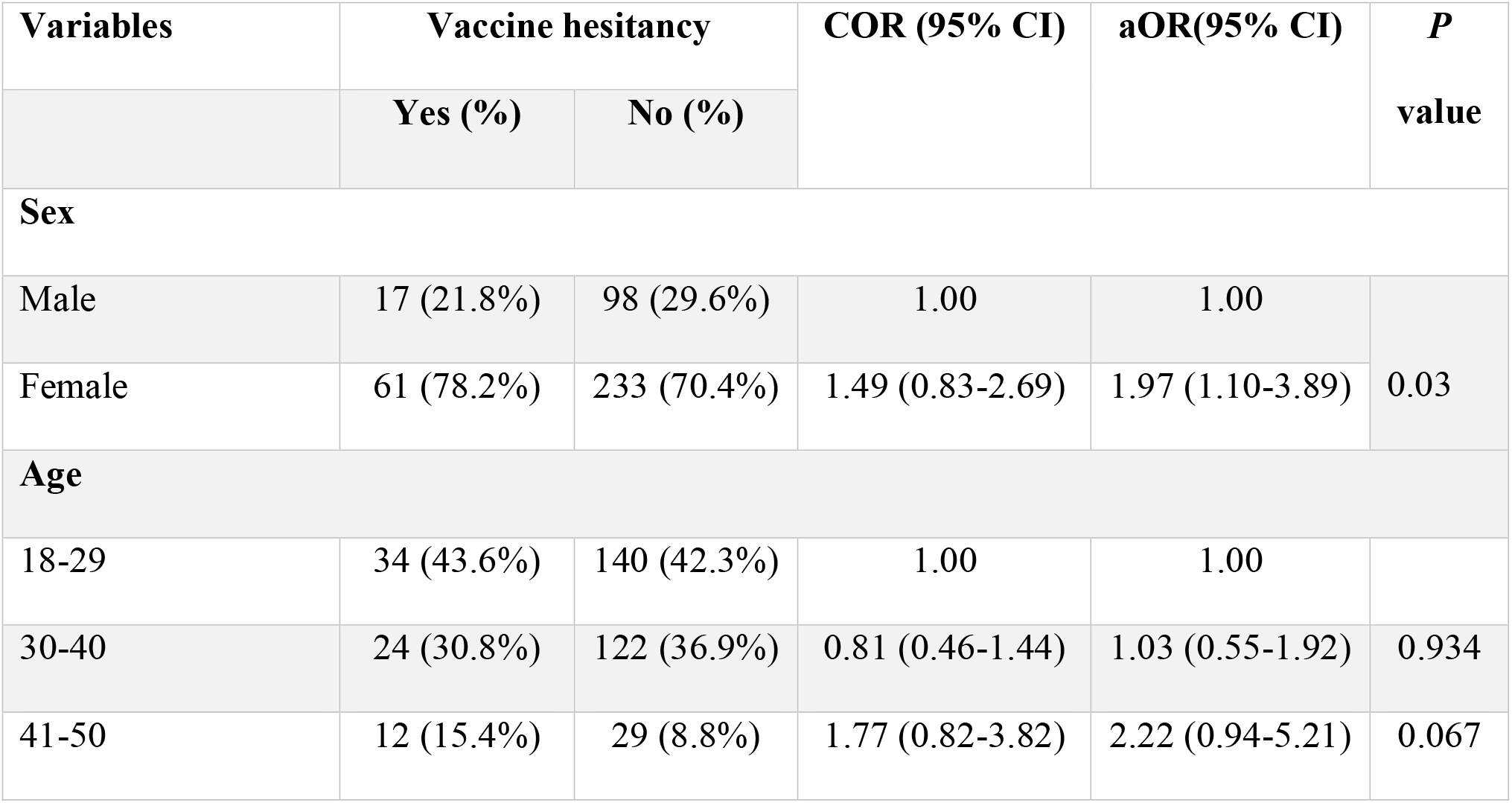
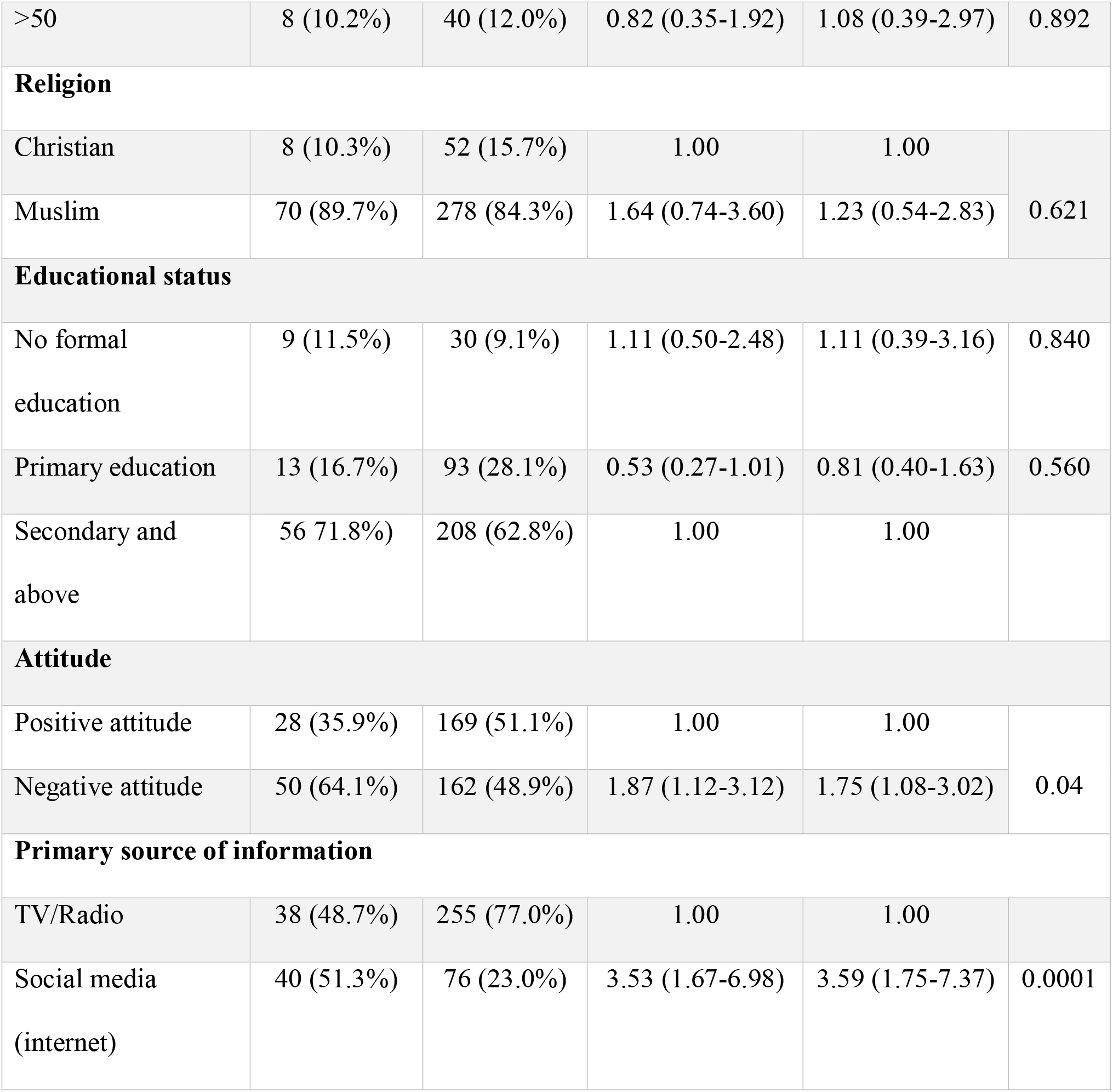
*Factors associated with COVID-19 vaccine hesitancy in Addis Ababa, Ethiopia, 2021*

It was found that the odds of vaccine hesitancy was 1.97 times (aOR=1.97; 95% CI: 1.10 - 3.89, p=0.03) higher among female participants as compared to male participants. The odds of vaccine hesitancy was 1.75 (aOR=1.75; 95% CI:1.08 - 3.02, p=0.04) times higher in those participants who were found to have a negative attitude towards COVID-19 and its preventive measures as compared to those who had a positive attitude. Similarly, the odds of vaccine hesitancy was 3.6 times (aOR=3.59; 95% CI: 1.75 - 7.37, P <0.0001) higher among those participants that received their information from social media (internet) as compared to those who received information only from TV/radio.

## Discussion

For the COVID-19 battle, the population adherence to preventive measures is crucial; however, it is mainly affected by their KAP toward the disease [1]. The findings of this study showed that nearly half of the study participants demonstrated inadequate knowledge of COVID-19, indicating a great knowledge gap. This finding is higher than studies conducted in other parts of Ethiopia such as Arbaminch (23.5%) and Gedeo (39.5%), and other low income countries such as Ghana (34.9%), and Malaysia (22.7%) [14-17]. The discrepancies might be due to differences in the community awareness creation through mass media and social media. Further, in our study, more than half the participants had negative attitude towards COVID-19 and its preventive measures, which is higher than the finding of a study conducted in Southern Ethiopia [15, 18] and lower than study done among Dessie and Kombolcha town residents in Ethiopia [19]. The discrepancy in the findings may be due to differences in the study period. The later studies were conducted earlier in the pandemic when the declaration and enforcement of state of emergency and other measures were still in place. Our findings show a significant decrease in the community’s attitude towards COVID-19 and its prevention measures which can lead people to become discouraged to consistently adhere to the measures set forth by the government and the world health organization. These findings of the study has an implication on the public health and underscore the need for urgent concerted efforts to promote the knowledge of the general public in Ethiopia towards COVID-19 preventive measures. If the current trend evidenced by this study continues in Ethiopia, COVID-19 will pose a devastating outcome on the medical, financial and social aspect of citizens besides the potential for new strains of disease developing.

As COVID-19 continues to ravage the world, vaccination offers the most reliable hope for a permanent solution to controlling the pandemic. However, a vaccine must be accepted and used by a large majority of the population to create herd immunity [20]. The findings of this study showed that about one out of five participants are not willing to receive COVID-19 vaccine when it is available, which is higher than the findings reported from developed countries such as UK (3%) [9, 21, 22]. The discrepancies might be due to insufficient knowledge about the vaccine and difference in the perception of the seriousness of the pandemic. This implies that if the doubts and fears of the majority regarding the vaccine are not addressed properly, we may not be able to attain herd immunity. Surprisingly, the finding of this study was lower than a study conducted in the US (31%) and Nigeria (80%) [13, 20]. This might be due to difference in access to wide variety of conspiracy theories and doubts via internet.

Consistent to the study conducted in China [23], vaccine hesitancy was more likely among females as compared to males in our study. This could be due to higher exposure of males for different media as compared to females in Ethiopia. In the present study, increased likelihood of vaccine hesitancy was also indicated among those with negative attitude towards COVID-19 and its preventive measures. The qualitative aspects of this study also found that those participants who would not take the vaccine stated one of their reason to be their lack of implicit trust in the government and in health professionals. Thus, this lack of confidence in the government exhibited by 41.8% of our participants may be a potential hurdle we might face after the arrival of the vaccine to Ethiopia.

In our study, those participants who received their information from social media (internet) were more likely to have vaccine hesitancy as compared to those who got their information only from TV/radio. This finding of the study is in line with a study conducted to assess health protective behaviors and conspiracy theories during the pandemic found that there was significant association between holding a conspiracy belief and checking social media for news of COVID-19 [24]. As a result, this finding is justified by our findings on both the quantitative and qualitative aspects of our study which found that the majority of the reasons given for hesitancy towards the vaccine were the belief in the conspiracy theories. Thus, the spared of these conspiracy theories is a potential issue that can cause problems when vaccine distribution starts in Ethiopia. Particularly, if these conspiracy theories start getting a wider audience thus there may be a need to act in haste and find a solution before this issue worsens.

This study is the first community based study to assess the Ethiopian community’s perception towards COVID-19 vaccine and its level of acceptance. We employed a mixed methods design which enables us to make the deep understanding of the issue. However, the study might be limited due to the recall bias and social desirability bias during the data collection. In addition to this, our sample over represents female population because the majority of the study participants that were found at home during data collection time were housewives. Therefore, generalization of the study results need to be cautious.

## Conclusion

A considerable proportion of the people have concerns of the COVID-19 vaccine and unwilling to accept once it is available. Several conspiracy theories were put forth to justify their stance and this was mainly due to the misconceptions distributed from the use of social media as source of information about the vaccine. Overall, providing the community with health education and consistent government efforts in uphold the prevention measures are of paramount importance to tackle this pandemic.

## Data Availability

Data are available upon reasonable request from the corresponding author.

## List of Abbreviations

aOR: Adjusted OddsRatio
CI: Confidence Interval
COR: Crude Odds Ratio
SARS: Severe Acute Respiratory Syndrome
UK: United Kingdom
US: United States

## Availability of data and material

Data are available upon reasonable request from the corresponding author.

## Authors Contributions

All the authors have substantially involved in the design of the study, collection of data, visualization of data, analysis of the data, and interpretation of the results, and drafted the initial manuscript and approved the final manuscript. All authors have read and approved the manuscript.

## Acknowledgement

The authors would like to thank Myungsung Medical College for providing fund to conduct this study. The authors are grateful to the study participants for their contributions.

