## Supplementary material for "COVID-19 Vaccine hesitancy in Addis Ababa, Ethiopia: A mixed-methods study": (Supplementary Table 1)

Table: Knowledge towards COVID 19 among residents of Addis Ababa, Ethiopia, 2021

|  | Questions | Correct count  N (%) | Incorrect count  N(%) |
| --- | --- | --- | --- |
| K1 | Transmission modes of COVID-19 |  |  |
|  | Is it transmitted by respiratory droplets? | 401 (98.0%) | 8 (2.0%) |
|  | Is it airborne? | 323 (79.0%) | 86 (21.0%) |
|  | Is it transmitted feco-orally? | 111 (27.1%) | 298 (72.9%) |
|  | Is it transmitted by blood? | 342 (83.6%) | 67 (16.4%) |
|  | Is it transmitted by touching contaminated surfaces? | 397 (97.1%) | 12 (2.9%) |
|  | Is it transmitted by consuming contaminated food? | 212 (51.8%) | 197 (48.2%) |
|  | Is it transmitted by contact with an infected person? | 390 (95.4%) | 19 (4.6%) |
|  | Is it transmitted by touching? | 401(98.0%) | 8 (2.0%) |
|  | Is it transmitted by Breast milk? | 343 (83.9%) | 66 (16.1%) |
|  | Is it transmitted from mother to child? | 337 (82.4%) | 72 (17.6%) |
| K2 | Symptoms of COVID-19 |  |  |
|  | Is fever a symptom? | 400 (97.8%) | 9 (2.2%) |
|  | Is myalgia a symptom? | 209 (51.1%) | 200 (48.9%) |
|  | Is fatigue a symptom? | 346 (84.6%) | 63 (15.4%) |
|  | Is diarrhoea a symptom? | 141 (34.5%) | 268 (65.5%) |
|  | Is sneezing a symptom? | 55 (13.4%) | 354 (86.6%) |
|  | Is losing your sense of smell a symptom? | 188 (46.0%) | 221 (54.0%) |
|  | Is vomiting a symptom? | 166 (40.6%) | 243 (59.4%) |
|  | Is rhinorrhoea a symptom? | 214 (52.3%) | 195 (47.7%) |
|  | Is SOB a symptom? | 340 (83.1%) | 69 (16.9%) |
|  | Is cough a symptom? | 392 (95.8%) | 17 (4.2%) |
|  | Is loss of taste a symptom? | 199 (48.7%) | 210 (51.3%) |
|  | Is stuffy nose a symptom? | 169 (41.3%) | 240 (58.7%) |
|  | Is conjunctivitis a symptom? | 77 (18.8%) | 331 (80.9%) |
|  | Is skin rash a symptom? | 34 (8.3%) | 375 (91.7%) |
|  | Can a covid-19 patient be asymptomatic? | 324 (79.2%) | 85 (20.8%) |
| K3 | Are asymptomatic patients capable of transmitting the disease? | 322 (78.7%) | 87 (21.3%) |
| K4 | Do old people have likelihood of developing severe disease? | 402 (98.3%) | 7 (1.7%) |
|  | Do pregnant women have likelihood of developing severe disease? | 259 (63.3%) | 150 (36.7%) |
|  | Do children have likelihood of developing severe disease? | 282 (68.9%) | 127 (31.1%) |
|  | Do smokers have likelihood of developing severe disease? | 360 (88.0%) | 49 (12.0%) |
|  | Do people with co-morbid conditions have likelihood of developing severe disease? | 396 (96.8%) | 13 (3.2%) |
|  | Do obese people have likelihood of developing severe disease? | 183 (44.7%) | 226 (55.3%) |
| K5 | Are you aware that hand washing is one of the primary methods of preventing COVID-19 infection? | 408 (99.8%) | 1 (0.2%) |
| K6 | Which is the most preferable method of hand washing to prevent COVID-19? | 374 (91.4%) | 35 (8.6%) |
| K7 | What is the recommended minimum duration of handwashing? | 242 (59.2%) | 167 (40.8%) |
| K8 | Do you think the use of face masks can prevent COVID-19 transmission? | 405 (99.0%) | 4 (1.0%) |
| K9 | Do you think double mask use is a more effective prevention method? | 203 (49.6%) | 206 (50.4%) |
| K10 | What’s the recommended minimum distance to maintain for adequate social distancing? | 268 (65.5%) | 141 (34.5%) |
| K11 | In order to prevent spread, do you think individuals should avoid going to crowded places and taking public transportation? | 383 (93.6%) | 26 (6.4%) |
| K12 | Do you think you should avoid social distancing provided that you’re wearing a mask? | 331 (80.9%) | 78 (19.1%) |
| K13 | Do you think you should avoid shaking hands and hugging when greeting people? | 401 (98.0%) | 8 (2.0%) |
| K14 | Provided that your family member is COVID-19 positive, would you put yourself in self-quarantine? | 381 (93.2%) | 28 (6.8%) |
| K15 | How long should people who were in contact with COVID-19 positive individuals self-isolate? | 281 (68.7%) | 128(1.3%) |
