## Supplementary material for "COVID-19 Vaccine hesitancy in Addis Ababa, Ethiopia: A mixed-methods study": (Supplementary Table 2)

Table: Attitude of the participants towards COVID-19 and its preventive measures

|  | Questions | Yes | No |
| --- | --- | --- | --- |
|  |  | Count (%) | Count (%) |
| A1 | Do you agree that COVID-19 will be successfully controlled? | 338 (82.6%) | 71 (17.4%) |
| A2 | I have concern of being infected with COVID-19 | 287 (70.2%) | 122 (29.8%) |
| A3 | Do you have confidence that Ethiopia will win the battle against COVID-19? | 281 (68.7%) | 128 (31.3%) |
| A4 | Is the Ethiopian government handling the COVID-19 health crisis well? | 238 (58.2%) | 171 (41.8%) |
| A5 | Do you think that wearing a face mask will effectively prevent COVID-19? | 401 (98.0%) | 7 (1.7%) |
| A6 | Do you think that adequate social distancing will effectively prevent COVID-19? | 405 (99.0%) | 4 (1.0%) |
| A7 | Do you think washing hands with soap and water helps to prevent COVID-19? | 405 (99.0%) | 4 (1.0%) |
| A8 | Would you be willing to tell people if you were having COVID-19 symptoms? | 401 (98.0%) | 8 (2.0%) |
| A9 | Would you inform the health authorities if a family member exhibits the symptoms? | 407 (99.5%) | 2 (0.5%) |
| A10 | Do you think traditional medicine can prevent or treat COVID-19? | 122 (29.8%) | 287 (70.2%) |
| A11 | Do you think COVID-19 doesn’t affect youngsters? | 50 (12.2%) | 359 (87.8%) |
